# Antiandrogen Flutamide-Induced Restoration of miR-449 Expression Mitigates Functional Biomarkers Associated with Ovarian Cancer Risk

**DOI:** 10.1101/2024.02.26.24303311

**Authors:** Xia Wang, Ho-Hyung Woo, Michele Wei, Steven Gibson, Mitzi Miranda, Demaretta Rush, Janiel Cragun, Wenxin Zheng, Guang Yao, Setsuko K Chambers

## Abstract

**Background:** The involvement of the androgen and androgen receptor (AR) pathway in the development of epithelial ovarian cancer is increasingly recognized. However, the specific mechanisms by which anti-androgen agents, such as flutamide, may prevent ovarian cancer and their efficacy remain unknown. We examined the effects of flutamide on the miRNA expression profile found in women at high risk (HR) for ovarian cancer.

**Methods:** Ovarian and tubal tissues, free from ovarian, tubal, peritoneal cancers, and serous tubal intraepithelial carcinoma (STIC), were collected from untreated and flutamide-treated HR women. Low-risk (LR) women served as controls. Transcriptomic miRNA sequencing was performed on these 3 sample cohorts. The miRNAs that showed the most notable differential expression were subjected to functional assays in primary ovarian epithelial cells and ovarian cancer cells.

**Results:** Flutamide treatment demonstrated a normalization effect on diminished miRNA levels in HR tissues compared to LR tissues. Particularly, the miR-449 family was significantly upregulated in HR ovarian tissues following flutamide treatment, reaching levels comparable to those in LR tissues. MiR-449a and miR-449b-5p, members of the miR-449 family, were computationally predicted to target the mRNAs of AR and colony-stimulating factor 1 receptor (CSF1R, also known as *c-fms)*, both of which are known contributors to ovarian cancer progression, with emerging evidence also supporting their roles in ovarian cancer initiation. These findings were experimentally validated in primary ovarian epithelial cells and ovarian cancer cell lines (SKOV3 and Hey): flutamide treatment resulted in elevated levels of miR-449a and miR-449b-5p, and introducing mimics of these miRNAs reduced the mRNA and protein levels of CSF1R and AR. Furthermore, introducing miR-449a and miR-449b-5p mimics showed inhibitory effects on the migration and proliferation of ovarian cancer cells.

**Conclusion:** Flutamide treatment restored the reduced expression of miR-449a and miR-449b-5p in HR tissues, thereby decreasing the expression of CSF1R and AR, functional biomarkers associated with an increased risk of ovarian cancer. In addition to the known direct binding of flutamide to the AR, we found that flutamide also suppresses AR expression via miR-449a and miR-449b-5p upregulation, revealing a novel dual-inhibitory mechanism on the AR pathway. Taken together, our study highlights mechanisms supporting the chemopreventive potential of flutamide in ovarian cancer, particularly in HR patients with reduced miR-449 expression.

## Introduction

Post-transcriptional RNA regulation is pivotal in modulating gene expression, with profound impacts on various cellular activities. As such, it has attracted significant interest in cancer research. Reflecting this focus, our studies have examined related regulatory mechanisms mediated by miRNAs, RNA-binding proteins, RNA epigenetics, and convergent gene pairs leading to antisense RNA regulation in breast and ovarian cancers [1–4].

Using prospectively collected biospecimens from predefined cohorts, we leveraged “bedside-to-bench” reverse translational research in hypothesis-driven, investigator-initiated trials (IIT). Central to our research has been the pursuit of early detection and prevention for women who are at HR for developing ovarian cancer [5–8]. Consequently, we initiated a phase 2 study (flutamide IIT NCT00699907) to assess the role of the anti-androgen flutamide in modulating gene expression and influencing potential biomarkers in HR women. This initiative was informed by our previous in vitro and in vivo studies that underscored the potential role of androgen signaling in ovarian cancer development [9]. These findings steered us towards evaluating anti-androgens, such as flutamide, as a potential preventive measure for HR women. Furthering this line of inquiry, we also conducted a parallel study to assess biomarkers of ovarian cancer risk in HR versus LR women [10].

In the initial targeted analysis of the flutamide IIT, immunohistochemistry revealed the overexpression of CSF1/CSF1R (macrophage colony-stimulating factor and its receptor) and ErbB4 proteins in tissues from HR women compared to LR women. Notably, this overexpression was attenuated in HR women who underwent flutamide treatment. Together, our findings underscore the potential involvement of cellular pathways such as CSF1/CSF1R and androgen receptor (AR) in the development of epithelial ovarian cancer and the response to flutamide treatment [9, 10].

In our ongoing Flutamide Correlative Study—a reverse translational research initiative—we seek to delineate global variations in miRNA profiles across three distinct cohorts: LR, HR, and flutamide-treated HR. Considering the extensive roles of miRNAs in governing RNA regulation and subsequent cellular processes, we hypothesize that miRNA profiling may differentiate HR and LR cohorts. We also postulate that flutamide treatment may, to some extent, correct the aberrant miRNA profile observed in HR individuals, potentially impacting functional biomarkers such as CSF1/CSF1R.

Specifically, the Flutamide Correlative Study aims to identify miRNAs differentially expressed between HR and LR women which regulate the dysregulated CSF1/CSF1R and AR pathways in HR patients. For this, we analyzed tissue samples from HR women, comparing them to both flutamide-treated HR and LR samples. Notably, our study excluded women diagnosed with ovarian or other peritoneal cancers. This exclusivity sets our research apart, as we focus solely on morphologically and pathologically normal ovary and fallopian tube samples from cancer-free women. In contrast, the existing literature to date on miRNA profiling in ovarian cancer primarily focuses on women already diagnosed with the disease [11–15].

In this study, our miRNA-sequencing results highlighted that flutamide treatment leads to a significant upregulation of the miR-449 cluster in the HR cohort, effectively restoring its expression to levels seen in LR women. Subsequent in vitro analyses in ovarian cells corroborated that miR-449a and miR-449b-5p reduce the expression of CSF1R and AR. Consistent with CSF1R and AR pathways being known contributors to ovarian cancer progression and initiation and being significantly elevated in HR women, we observed that miR-449a and miR-449b-5p reduced the migration and proliferation of ovarian cancer cells. Remarkably, our study unveiled a novel mechanism wherein the anti-androgen flutamide suppresses AR expression by upregulating miR-449, complementing its well-known role as a direct-binding AR inhibitor. Collectively, these data validate our miRNA profiling approach in the ongoing Flutamide Correlative Study and strengthen the basis for flutamide as a chemopreventive approach in HR women with downregulated miR-449 expression.

## Methods

### Definition of High-Risk (HR)

HR women were identified based on the following criteria in this study: carrying a BRCA1 or BRCA2 mutation, a Lynch Syndrome mutation, and/or having a family history characterized by one or more first-degree relatives with epithelial ovarian cancer, one or more cases of breast cancer diagnosed at age 40 or younger, more than one case of breast cancer diagnosed at age 50 or younger, instances of male breast cancer, and/or a family history of breast/ovarian cancer. Among the HR group, there was a higher prevalence of BRCA2 mutations compared to BRCA1, representative of our HR clinic population [6]. Low-risk (LR) patients did not meet any of these HR criteria. A personal history of breast cancer was present in both HR and LR populations [9, 10].

### Clinical matching of patient cohorts

This report is a validation of a notable finding from the initial miRNA sequencing data obtained in our overarching and ongoing Flutamide Correlative Study. In the study, we designed a comparison across 3 cohorts (HR, LR, and flutamide-treated HR). The flutamide-treated HR cohort consisted of 12 available patients, and for a balanced comparison, similar numbers of patients from the HR (n=12) and LR (n=13) groups were selected, considering matching clinical parameters and tissue sample collection.

The HR and LR cohorts were chosen to match as closely as possible to the flutamide treated-HR cohort and to each other in terms of menopausal status (post-menopausal vs. peri- or pre-menopausal) and Body Mass Index (BMI). There were 9 of 12 (75%) peri-/pre-menopausal patients in the HR cohort and 9 of 13 (69%) such patients in the LR cohort, compared to 9 of 12 (75%) in the flutamide-treated HR cohort. The median BMI (range) in the LR cohort was 27.9 kg/m^2^ (19.9-36.0), compared to 27.1 kg/m^2^ (21.0-36.9) in the HR cohort and 25.5 kg/m^2^ (19.3-45.0) in the flutamide-treated HR cohort. Additionally, the HR and flutamide-treated HR cohorts were matched for BRCA1/2 status, with 9 of 12 (75%) patients in each cohort carrying either a BRCA1 or BRCA2 germline mutation.

### Sample Preparation and small RNA extraction

Deidentified fixed fallopian tube and ovary tissues were obtained following patient consent, as outlined in the section on clinical matching of patient cohorts. Both ovarian and fallopian tube samples were included in this study, considering that the fallopian tubal epithelium has been proposed as a potential origin of ovarian epithelial carcinoma. None of the fallopian tubes included in this study had tubal dysplasia or serous tubal intraepithelial carcinoma (STIC), as determined by our Gynecologic Oncology pathologists.

Tissue sections (10 µm) were prepared from each ovarian and tubal sample. In instances where ovary and tube samples were found embedded within the same block, they were meticulously separated and re-embedded. We note that ovarian tissue blocks from 2 of 13 LR cases were unable to be retrieved from archives, with 2 of 12 fallopian tube blocks unavailable from the flutamide-treated HR cases.

Small RNAs were extracted from an average of 10 sections per sample using the miRNeasy FFPE kit (Qiagen). Quality tests, including qRT-PCR and gel electrophoresis, were used to confirm successful miRNA isolation without contamination. Our miRNA analysis included a total of 12 HR, 12 flutamide-treated HR, and 11 LR ovarian samples, along with a total of 12 HR, 10 flutamide-treated HR, and 13 LR fallopian tube samples.

### miRNA-sequencing and analysis

Small RNA libraries were constructed and sequenced following a standardized procedure at GENEWIZ (South Plainfield, NJ). Libraries were prepared using the TruSeq Small RNA library Prep Kit (Illumina, San Diego, CA), validated on the Agilent TapeStation 4200 (Agilent Technologies, Palo Alto, CA, USA), and quantified using a Qubit 2.0 Fluorometer (Invitrogen, Carlsbad, CA). Sequencing was performed on an Illumina HiSeq instrument in a 2×150 bp paired-end configuration. The sequencing reads have been deposited in the Gene Expression Omnibus (GEO) database (GSE252170).

For data processing, sequencing adapters and low-quality bases were trimmed using Cutadapt [27]. Reads with 18-26 nt in length were kept, and the paired-end reads were merged using fastq-join [28]. Reads mapping and miRNA quantification were performed using miRMaster with the default parameter settings [29, 30]. Differential expression analysis was performed using DESeq2 [31] with a negative binomial generalized linear model. A miRNA was considered differentially expressed between two sample groups if it exhibited > 2-fold difference in mean expression with a p-value < 0.05. Potential mRNA targets of miRNAs were identified using the miRDB database [16], based on MirTarget predictions derived from extensive miRNA-target interaction analyses [32].

### Cell culture and flutamide treatment

SKOV3 and Hey human ovarian cancer cells were cultured in Dulbecco’s Modified Eagle/F12 Ham’s medium (DMEM/F12) with 10% NuSerum (Corning). Primary ovarian epithelial cells (Cell Biologics, Chicago, IL) were cultured in DMEM/F12 with hEGF (400 pg/ml) and 10% NuSerum. The HR or LR origin of these primary ovarian epithelial cells is not known. The limited information (donor age between 56-60 years) may indicate a LR rather than HR origin, as HR women would typically undergo removal of normal ovaries at an earlier age. All cells were incubated at 37°C with 5% CO_2_. For flutamide treatment, cells were cultured in DMEM/F12 supplemented with 1% NuSerum for 24 hours, followed by a 24-hour treatment with flutamide (5 µM) in the same medium before being harvested for analysis.

### Quantitative reverse transcription PCR (qRT-PCR)

Total RNA was extracted with Trizol (Invitrogen) and treated with RNase-free DNase (Promega, USA). For miRNA quantification, stem-loop qRT-PCR was used to enhance amplification specificity [33]. Following cDNA synthesis of miRNAs and 5S rRNA internal control, quantitative PCR (qPCR) was conducted using an Eppendorf Realplex2 instrument. The thermal cycling conditions included a 10-minute initial denaturation at 95 °C, followed by 40 cycles of 15 seconds at 95 °C and 1 minute at 60 °C. Final miRNA expression values were calculated using the ΔΔCt method [34]. Similarly, for mRNA quantification, CSF1R and AR transcripts were reverse transcribed by pdN6 random hexamers and quantified with qPCR, with GAPDH transcript used as an internal control. All qRT-PCR primer sequences are detailed in Table 1.

**Table 1.**
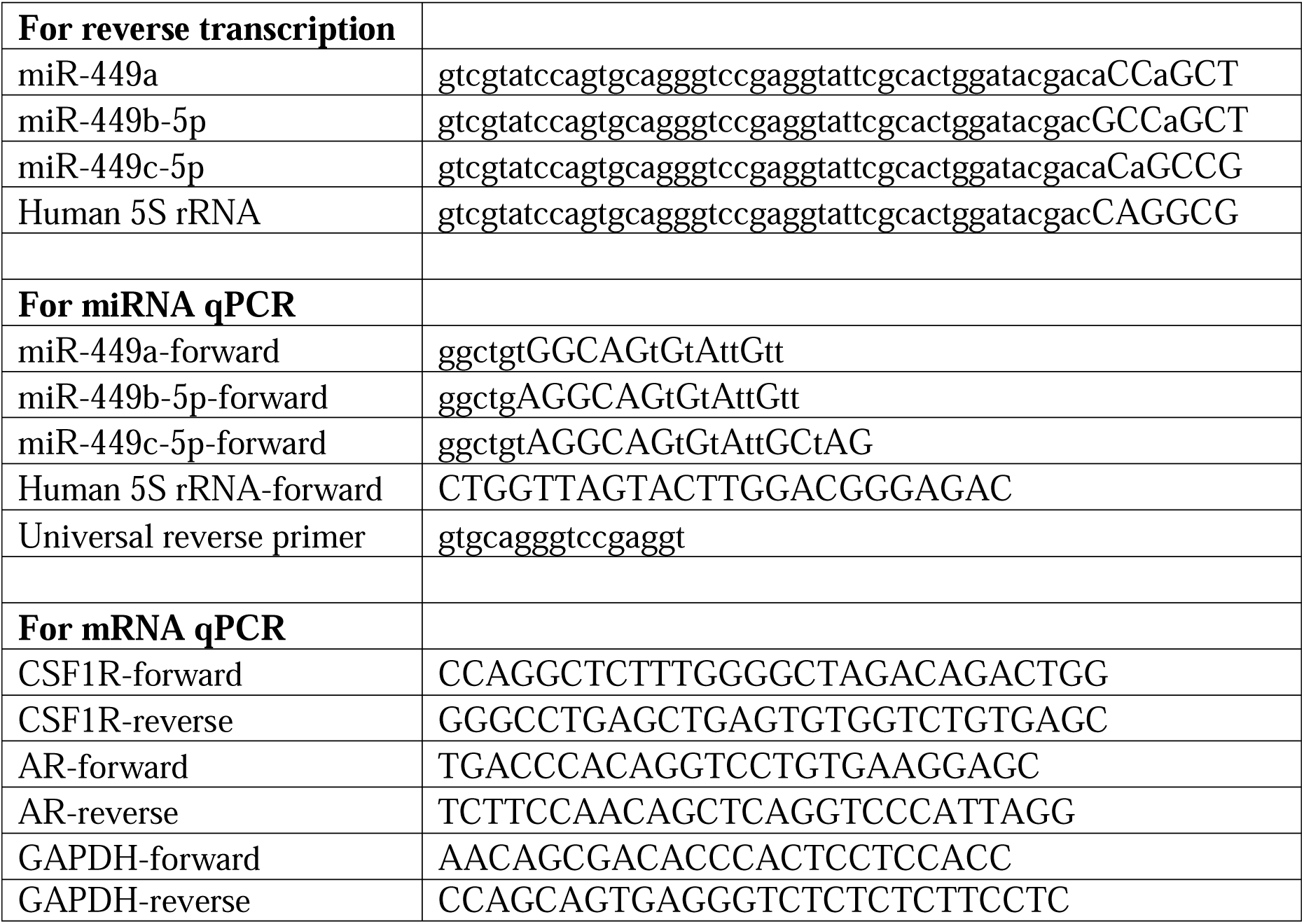
Primers used for qRT-PCR.

### Transient transfection with miRNA mimics

Cells were plated on 6-well plates one day prior to transfection. Mimics of miR-449a and miR-449b-5p were purchased from Sigma (MISSION microRNA Mimics). Transfection of miRNA mimics was performed according to the manufacturer’s instructions, alongside MISSION miRNA negative control.

### Immunoblotting

Cytoplasmic lysates (140 μg), isolated with NE-Per (Thermofisher, USA), were subjected to SDS-PAGE and electroblotted onto a PVDF membrane. The membranes were incubated with mouse monoclonal anti-c-fms (sc-46662, Santa Cruz Biotech, USA), anti-AR (sc-7305, Santa Cruz Biotech, USA), and anti-Pan Actin (ACTN05, NeoMarkers, Fremont, CA) antibodies. Following washes with PBS-T (PBS with 0.1% Tween-20) and incubation with anti-mouse (HRP)-conjugated secondary antibody, proteins were detected using SuperSignal chemiluminescence (Pierce).

### Directed Migration and cell proliferation assays

Cell migration was quantified using Membrane Migration Culture Systems (Sigma, USA) as previously described [2]. Briefly, fibronectin (25 μg/ml, Fisher) was added as a chemoattractant in the lower wells of a 24-well plate, above which uncoated filters (8-μm pore) were placed. Cells (5×10^4^ per well) were seeded onto the uncoated filters and incubated for 20 hours. Cell proliferation was assessed using the WST-1 assay reagent (Cell Biolabs). Cells (1×10^3^ per well seeded in a 96-well plate) were stained with WST-1 every 24 hours for 3 days.

### Statistical analysis

Unless otherwise noted, data are presented as mean ± s.e.m. derived from experiments conducted in triplicate, analyzed using R with a one-sided unpaired t-test, considering a p-value < 0.05 indicative of statistical significance.

## Results

### miR-449 downregulation is associated with a high risk for ovarian cancer and can be reverted by flutamide treatment

In this study, we first conducted miRNA-sequencing profiling in the two ovarian cancer-related tissues, the ovary and fallopian tube, from HR and LR patients (high- and low-risk for ovarian cancer, respectively). HR group was further divided into two cohorts: those treated with flutamide (the “flutamide” cohort) and those not treated with flutamide (the “HR” cohort). The samples across the three cohorts (flutamide, HR, and LR) were matched for body mass index (BMI) and menopausal status. The HR and flutamide cohorts were defined for the high-risk status and balanced for germline BRCA1/2 conditions, as detailed in Methods.

Our analysis revealed notable differences in miRNA expression profiles between these cohorts. When comparing the flutamide cohort to the HR cohort, there was a significantly greater number of significantly up-regulated miRNAs than down-regulated (Suppl. Fig. 1A). A similar, though not as statistically significant, pattern was observed when comparing the LR cohort to the HR cohort (Suppl. Fig. 1B). These findings suggest a general trend where the HR status is associated with a lower miRNA expression profile compared to LR in both the ovary and tube.

Interestingly, this pattern appears to be reversed with flutamide treatment, indicating a potential regulatory role of flutamide in miRNA expression in these tissues.

Among these miRNA profiles (Suppl. Fig. 1), the most striking difference emerged from comparing the flutamide and HR cohorts in the ovary. Specifically, miR-449a, miR-449b-5p, and miR-449c-5p from the miR-449 family were identified among the most significantly up-regulated miRNAs (top right corner beyond the two red-dashed threshold lines, indicating fold change > 4 and p < 0.01, respectively; Fig. 1A). A similar trend of significant upregulation in miR-449a, miR-449b-5p, and miR-449c-5p was observed when comparing ovarian LR and HR cohorts (Suppl. Fig. 1B, left). That is, these miRNAs were highly expressed in LR, reduced in HR, and increased again with flutamide treatment in the ovary, a pattern confirmed by large effect size (Glass’s delta > 0.8) and statistical significance (p < 0.05; Fig. 1B, top). Interestingly, this expression pattern of the three miR-449 family members was also mirrored, albeit with a smaller effect size and statistical significance, in fallopian tube samples that were matched to the ovarian samples in our study (Fig. 1B, bottom).

**Figure 1.**
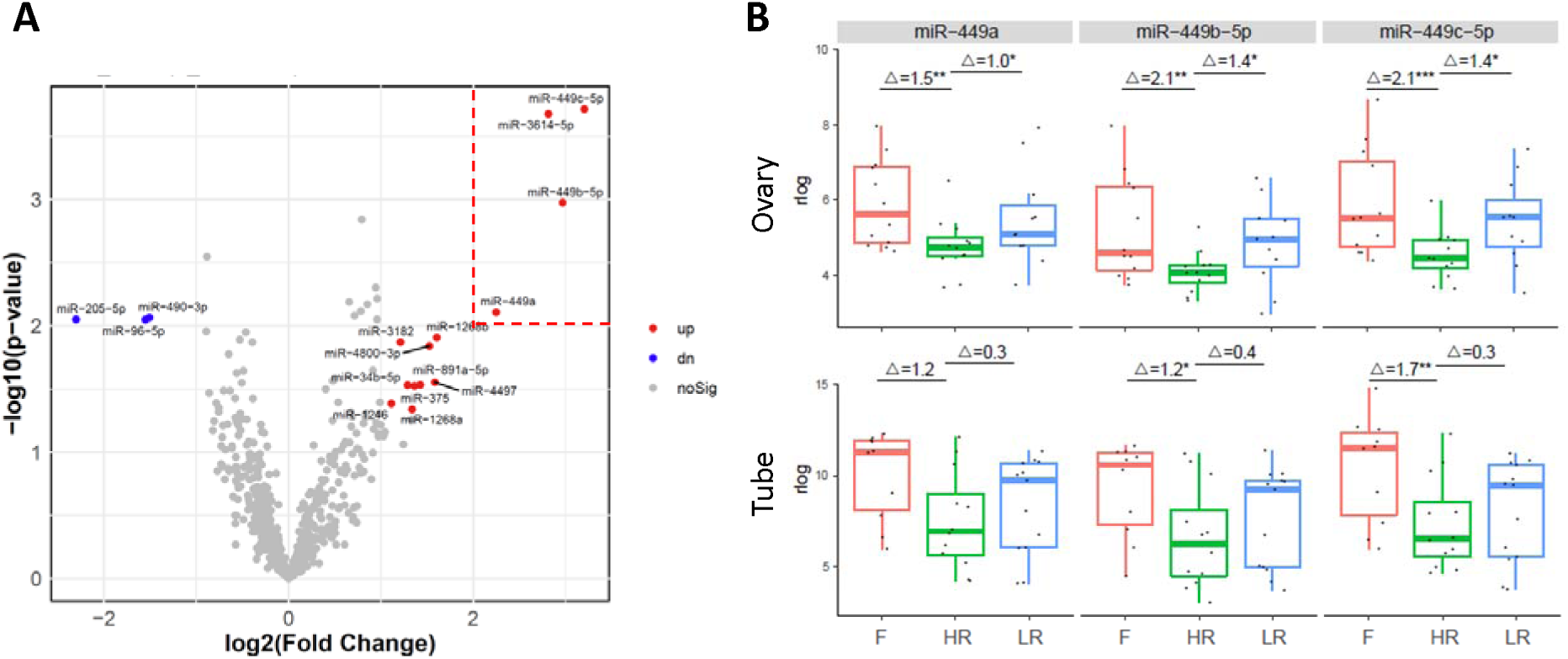
The miR-449 family is strongly associated with ovarian cancer HR and flutamide action. (**A**) Volcano plot of miRNA expression profile in flutamide vs. HR cohort from the ovary samples. Red and blue dots indicate miRNAs that are significantly up-regulated (fold change > 2, p < 0.05) and down-regulated (fold change < 0.5, p < 0.05), respectively. (**B**) Boxplot of the expression (in rlog normalized values, y-axis) of miR-449 family (miR-449a, left; miR-449b-5p, middle; miR-449c-5p, right) in the cohorts of flutamide (F), HR, and LR in the ovary (top) and fallopian tube (bottom). The effect size (Glass’s delta) and statistical significance (*, **, *** indicating p < 0.05, 0.01, 0.001, respectively) for each pairwise comparison based on DESeq2 differential analysis are shown at the top of each boxplot.

As an independent validation of the miRNA-seq results, we conducted qRT-PCR analysis on miR-449 expression using the original tissue samples. The qRT-PCR results corroborated the miRNA-seq data, showing not only a strong and significant correlation in miR-449 expression measurements across individual samples (Suppl. Fig. 2A) but also a consistent differential expression pattern between patient cohorts (high in LR, low in HR, and restored to high again following flutamide treatment) with even larger effect sizes than those observed in miRNA-seq (Suppl. Fig. 2B). These consistent observations, across both miRNA-seq and qRT-PCR analyses, suggest that in ovarian cancer-related tissues, the reduced expression of miR-449 family members correlates with the HR status for ovarian cancer, and flutamide treatment restores the expression of these miRNAs. Such miR-449-related patterns were particularly evident in the ovary, and thus the tissue focus of our subsequent analysis.

### miR-449a and miR-449b-5p suppression of CSF1R and AR expression as a link to flutamide action and ovarian cancer risk reduction

Next, we investigated whether and how miR-449a, miR-449b-5p, and miR-449c-5p may contribute mechanistically to the HR vs. LR status and flutamide action. Previously, we observed that i) androgen signaling promotes ovarian cancer development [9], and ii) CSF1/CSF1R and ErbB4 proteins were overexpressed in the ovarian tissues of HR women compared to LR, and this overexpression was reversed in those HR women treated with flutamide [9, 10]. Following these clues, we explored the computationally predicted mRNA targets of miRNAs associated with these HR-related proteins, based on the MirTarget tool in the miRDB database [16]. This analysis revealed that miR-449a and miR-449b-5p, but not miR-449c-5p, potentially target CSF1R and AR mRNAs (Suppl. Fig. 3), a finding that aligns with and potentially explains our previous observations (Fig. 2).

**Figure 2.**
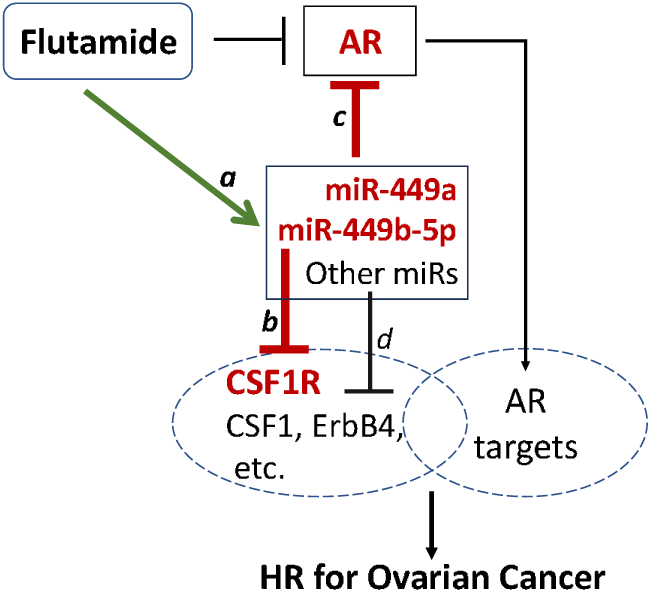
Working model of flutamide action on down-regulating oncogenic markers associated with ovarian cancer risk, such as CSF1R and AR, via the up-regulation of miR-449a and miR-449b-5p. *a*, Flutamide up-regulates miR-449a and miR-449b-5p (and other miRNAs); *b,* miR-449a and miR-449b-5p suppress CSF1R expression; *c*, miR-449a and miR-449b-5p suppress AR expression. *d*, Other flutamide-upregulated miRNAs may suppress CSF1, ErbB4, etc.

Specifically, our model posits the following sequence of events: first, given that miR-449a and miR-449b-5p may target and repress CSF1R expression (link *b*, Fig. 2; Suppl. Fig. 3), the downregulation of miR-449a and miR-449b-5p in HR relative to LR (Fig. 1B) could lead to the observed CSF1R overexpression in HR women [9]. Second, following flutamide treatment, miR-449a and miR-449b-5p are upregulated (link *a*, Fig. 2; Fig. 1), which could reverse the overexpressed CSF1R in HR women, consistent with the findings in [9]. Third, the computationally predicted inhibitory effects of miR-449a and miR-449b-5p on AR expression (link *c*, Fig. 2; Suppl. Fig. 3), if experimentally validated, could reveal a novel mechanistic link of flutamide action – that is, in addition to its established role as an AR antagonist by blocking the binding of androgens to AR, flutamide may also indirectly suppress AR expression by upregulating miR-449a and miR-449b-5p. In addition, we noticed that flutamide significantly upregulates several other miRNAs in HR ovarian and fallopian tube tissues (Suppl. Fig. 1A), which could target and suppress other HR markers such as CSF1, ErbB4, and possibly some AR targets (link *d*, Fig. 2). This current study primarily focuses on testing the upregulation of miR-449a and miR-449b-5p by flutamide in the ovary and their consequent effects on CSF1R and AR expression (links *a-c*, Fig. 2).

### Flutamide upregulates miR-449a and miR-449b-5p expression in both primary epithelial and malignant ovarian cells

In our miRNA-seq analysis, miR-449a and miR-449b-5p were significantly upregulated in ovarian samples from HR women treated with flutamide. Here, we first examined whether this relationship holds in experimental cell models before performing further mechanistic tests. To this end, we examined two ovarian cancer cell lines, SKOV3 and Hey, alongside primary ovarian epithelial cells. In each of the three ovarian cell models tested, we observed that both miR-449a and miR-449b-5p were upregulated upon flutamide treatment as measured by qRT-PCR (Fig. 3). This upregulation was statistically significant (p < 0.05) in the two ovarian cancer cell lines (SKO3 and Hey). In primary ovarian epithelial cells, the baseline expression of miR-449a and miR-449b-5p was significantly higher than that in SKO3 and Hey cells (Suppl. Fig. 4), which may account for their less pronounced, statistically non-significant, further upregulation observed post-flutamide treatment (Fig. 3).

**Figure 3.**
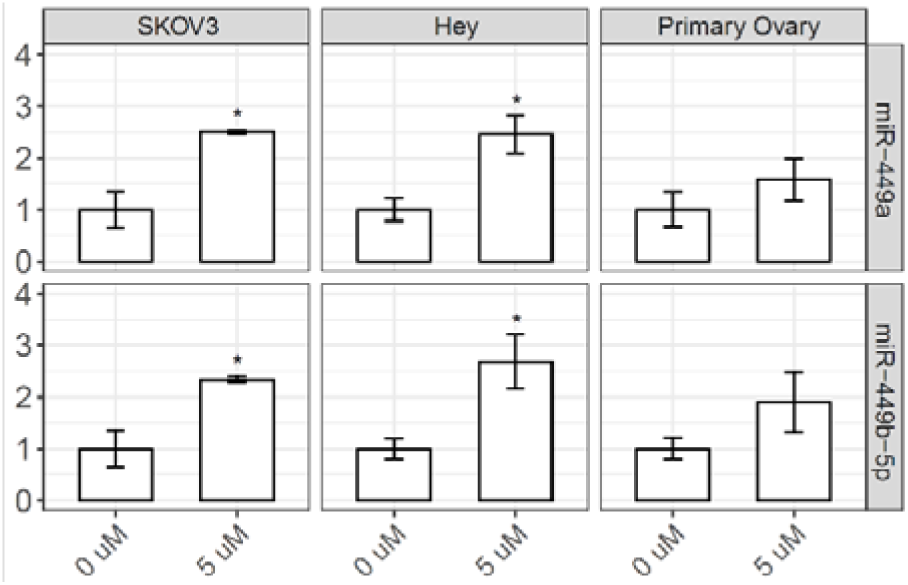
Flutamide up-regulates miR-449a and miR-449b-5p in both primary epithelial and malignant ovarian cells. Cells were treated with flutamide (5 µM) or vehicle control (0 µM) for 24 hours, and the expression of miR-449a (top) and miR-449b-5p (bottom) was determined by qRT-PCR (n=3, with the control expression set to 1). The statistical significance (if any) of each pairwise comparison to the vehicle control is indicated with * for p < 0.05. Error bar, s.e.m.

The lower baseline expression of miR-449a and miR-449b-5p in ovarian cancer cells, as compared to primary ovarian cells (Suppl. Fig. 4), are reminiscent of the pattern observed in our clinical samples, where these miRNAs were downregulated in HR ovarian samples relative to LR (Fig. 1B). Although as noted earlier, HR samples were morphologically and pathologically normal and cancer-free, these observations suggest a consistent trend of miR-449a and miR-449b-5p downregulation across the spectrum of ovarian cancer risk and progression.

### miR-449a and miR-449b-5p suppress CSF1R expression at both the mRNA and protein levels in ovarian cells

Next, we examined whether miR-449a and miR-449b-5p inhibit CSF1R expression as predicted computationally. To this end, we introduced the mimics of these miRNAs into each of the three ovarian cell models (Hey, SKOV3, and primary) and measured the corresponding CSF1R mRNA and protein levels. First, in Hey cells, we observed that mimics of miR-449a and miR-449b-5p, but not miR-449c-5p, showed pronounced effects in reducing the CSF1R expression at both the mRNA and protein levels compared to the mock control (Suppl. Fig. 5). These results are consistent with the computational prediction that miR-449a and miR-449b-5p, but not miR-449c-5p, target and suppress CSF1R (Suppl. Fig. 3). We therefore continued to focus on testing miR-449a and miR-449b-5p, not miR-449c-5p, in subsequent tests.

Similar results as in Hey cells were observed in SKOV3 ovarian cancer cells (top, Fig. 4) and the primary ovarian epithelial cells (bottom, Fig. 4). In both cell models, the introduction of miR-449a mimic led to a significant decrease in CSF1R mRNA and protein levels compared to the mock controls; the introduction of miR-449b-5p mimic resulted in a similar decrease in CSF1R mRNA and protein levels as with the miR-449a mimic, although the effect appeared less pronounced (and that on CSF1R protein in primary ovarian epithelial cells was insignificant), as seen in Fig. 4.

**Figure 4.**
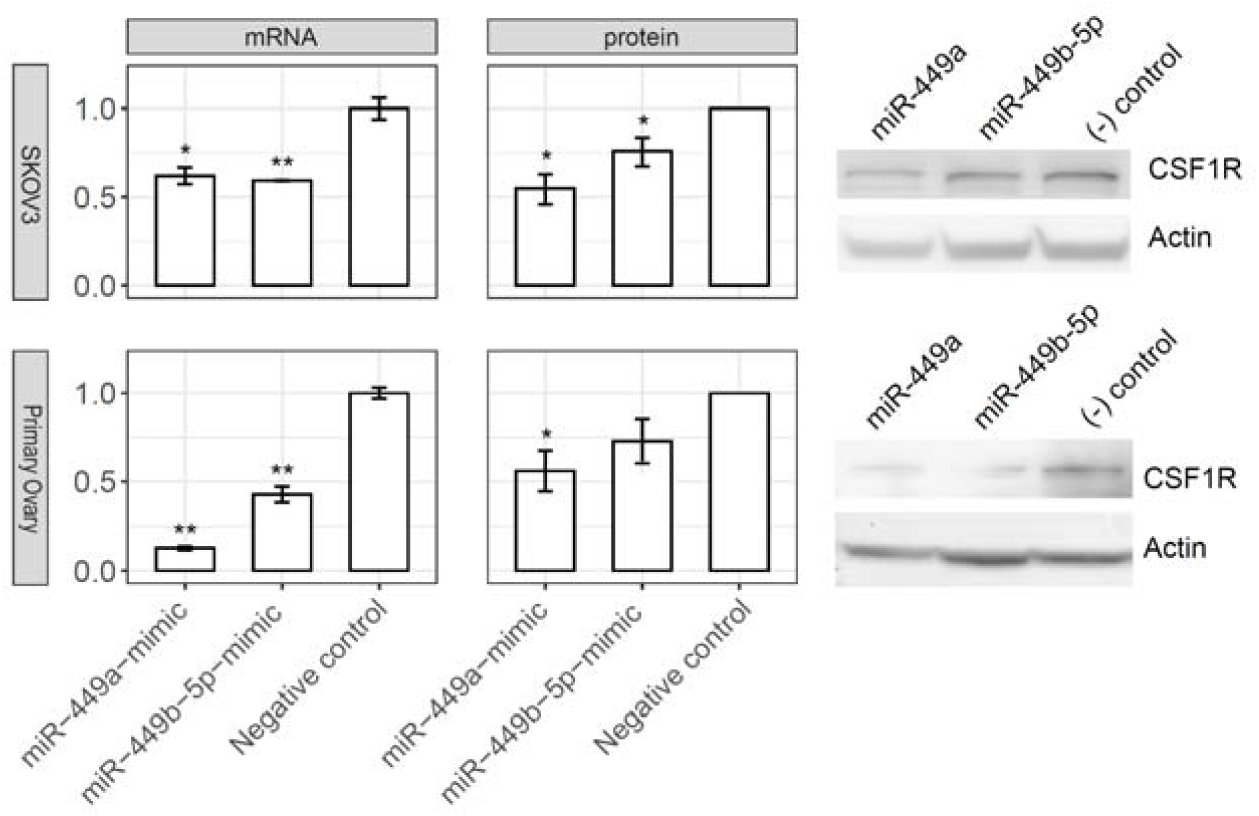
miR-449a and miR-449b-5p suppress CSF1R expression at both the mRNA and protein levels in ovarian cells. Upper panel, SKOV3 cells; Lower panel, Primary ovary cells. The mRNA level was determined by qRT-PCR (n=3) and the protein level by Western blot (n=2, with a representative blot shown to the right). The statistical significance (if any) of each pairwise comparison to the negative control is indicated with * and ** for p < 0.05 and 0.01, respectively. Error bar, s.e.m.

### miR-449a and miR-449b-5p repress AR expression at both the mRNA and protein levels in ovarian cancer cells

We subsequently examined whether miR-449a and miR-449b-5p inhibit AR expression as predicted computationally. To this end, we introduced the mimics of miR-449a and miR-449b-5p into the three ovarian cell lines (SKOV3, Hey, and Primary) and measured the corresponding AR mRNA and protein levels. We observed that miR-449a and miR-449b-5p mimics significantly reduced the AR mRNA and, consistently, AR protein levels, compared to the mock control in both ovarian cancer cell lines tested (SKOV3, upper panel, Fig. 5; Hey, Suppl. Fig. 6).

**Figure 5.**
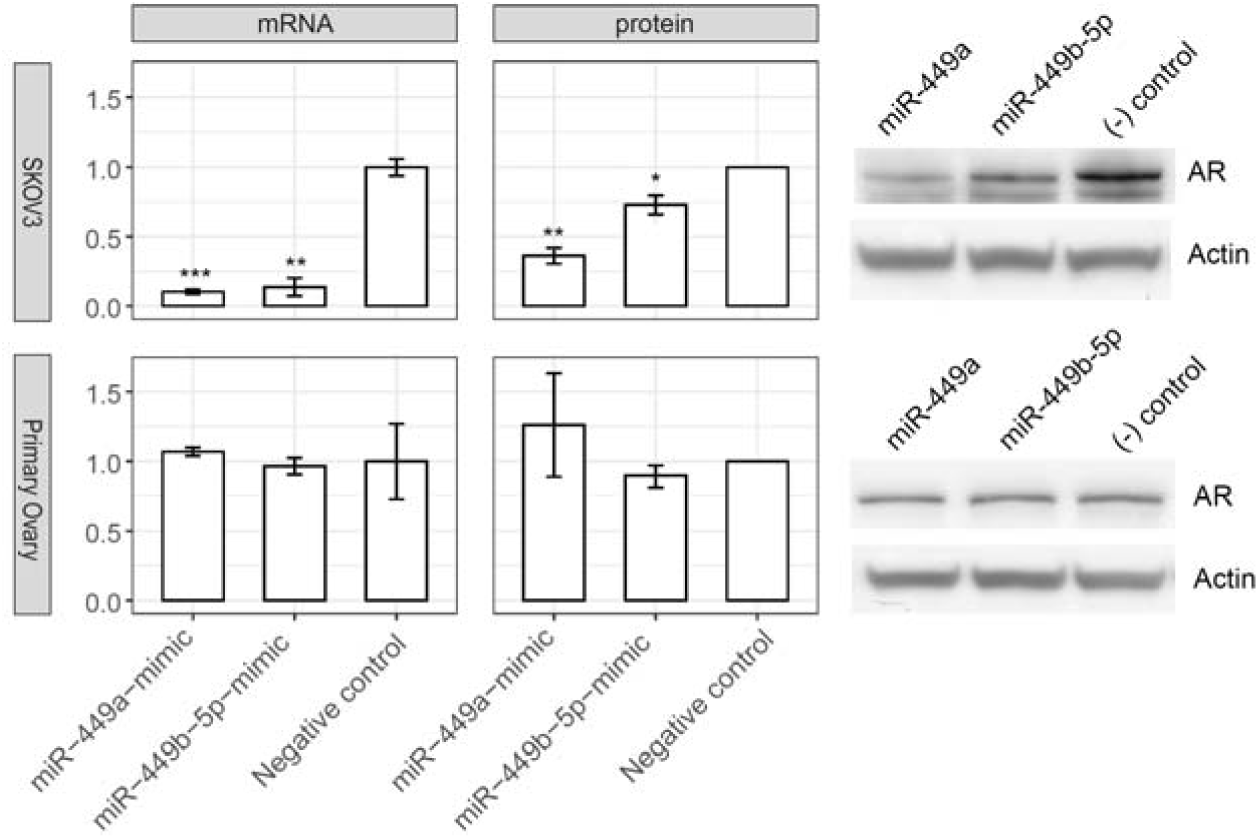
miR-449a and miR-449b-5p repress AR expression at both the mRNA and protein levels in ovarian cells. Upper panel, SKOV3 cells; Lower panel, Primary ovary cells. The mRNA level was determined by qRT-PCR (n=3) and the protein level by Western blot (n=2, with a representative blot shown on the right). The statistical significance (if any) of each pairwise comparison to the negative control is indicated with *, **, and *** for p < 0.05, 0.01, and 0.001, respectively. Error bar, s.e.m.

We did not observe a reduction in AR mRNA or protein levels following the introduction of miR-449a and miR-449b-5p mimics in primary ovarian epithelial cells (lower panel, Fig. 5). This discrepancy could be two-fold. *I*) The baseline levels of miR-449a and miR-449b-5p are significantly higher in primary ovarian epithelial cells than in Hey and SKOV3 ovarian cancer cells (Suppl. Fig. 4). Consequentially, the relative increase of these miRNAs via the introduced mimics is smaller in primary ovarian cells than in Hey and SKOV3 cells, likely leading to a less pronounced effect. *II*) The target scores of miR-449a and miR-449b-5p against AR (= 56 for both miRNAs) are lower than against CSF1R (= 73 and 68, respectively; Suppl. Fig. 3), indicating weaker interactions of these miRNAs with AR than with CSF1R mRNA. Consequently, the repressive effects on AR in primary ovarian cells are expected to be mild, considering both the small fold increase of miR-449a and miR-449b-5p levels via the introduced mimics and their potentially lower targeting efficiency to AR. This contrasts with the stronger effects of these miRNAs either on targeting AR in SKOV3 (Fig. 5, top) and Hey (Suppl. Fig. 6) cells, or on targeting CSF1R (Fig. 4, bottom) versus AR in primary cells, in line with our experimental results.

### miR-449a and miR-449b-5p inhibit the migration and proliferation of ovarian cancer cells

Since miR-449a and miR-449b-5p suppress CSF1R and AR expression in ovarian cancer cells (SKOV3 and Hey) and CSF1R and AR are associated with the risk of ovarian cancer development, we next tested whether these miRNAs could exert any noticeable effects on the behavior of ovarian cancer cells. To this end, we introduced miR-449a and miR-449b-5p mimics into SKOV3 and Hey cells. The endogenous baseline levels of these miRNAs are lower in these ovarian cancer cells than in primary ovarian epithelial cells (Suppl. Fig. 4), and thus, the effects of the introduced mimics may be better observed. We measured cell migration and proliferation in SKOV3 and Hey cells transfected with miR-449a and miR-449b-5p mimics versus the mock control. As seen in Fig. 6, both miR-449a and miR-449b-5p mimics induced a moderate, yet statistically significant, reduction in cell migration in both SKOV3 and Hey cells. In addition, introducing these miRNA mimics appeared to moderately reduce the proliferation of SKOV3 and Hey cells (Suppl. Fig. 7).

**Figure 6.**
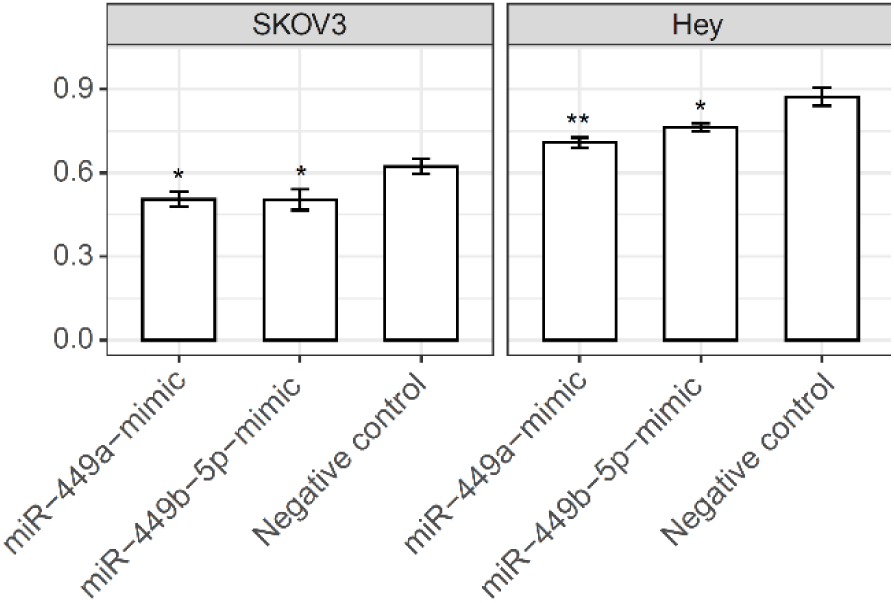
miR-449a and miR-449b-5p mimics decrease the directed motility of SKOV3 and Hey cells. Directed motility using fibronectin was performed as described in Methods (n=3). The statistical significance (if any) of each pairwise comparison to the negative control is indicated with * and ** for p < 0.05 and 0.01, respectively. Error bar, s.e.m.

## Discussion

In the present study, we performed transcriptomic miRNA profiling of ovarian and fallopian tube tissues from women at HR for ovarian cancer. A notable outcome of this analysis was the general trend of miRNA upregulation in response to anti-androgen flutamide treatment (Suppl. Fig. 1A). A similar upregulation pattern was also observed in tissues from women at LR for ovarian cancer when compared to their HR counterparts (Suppl. Fig. 1B). These results suggest that flutamide may exert a normalizing effect on the diminished miRNA levels in HR tissues, increasing them toward the levels observed in LR tissues. This finding is consistent with our earlier work, which demonstrated the role of flutamide in normalizing the expression of dysregulated gene markers such as CSF1, CSF1R, and ErbB4 in HR tissues [9, 10].

In our in vitro validations, we focused on miR-449a and miR-449b-5p, which are among the most significantly up-regulated miRNAs in response to flutamide treatment and computationally predicted to target and inhibit the expression of CSF1R and AR. We found that these miRNAs were significantly less expressed in SKOV3 and Hey ovarian cancer cell lines compared to the primary ovarian cell control (Suppl. Fig. 4), which aligns with their lower expression pattern in HR subjects related to LR patients (Fig. 1B). Upon further investigation in SKOV3 and Hey cells, we found that a) flutamide significantly upregulated the expression of miR-449a and miR-449b-5p, b) introducing mimics of miR-449a and miR-449b-5p downregulated CSF1R and AR expression at both mRNA and protein levels, and c) miR-449a and miR-449b-5p exhibited inhibitory effects on cell migration and proliferation (Figs. 3-6; Suppl. Figs. 5-7). In comparison, in primary ovarian epithelial cells, the upregulation of miR-449a and miR-449b-5p by flutamide was not statistically significant (Fig. 3), and miR-449a and miR-449b-5p mimics downregulated the expression of CSF1R (Fig. 4 bottom) but not AR (Fig. 5 bottom). These results are in line with the higher basal expression of these miRNAs in primary ovarian epithelial cells than in ovarian cancer cells (Suppl. Fig. 4) and their potentially higher targeting efficiency on CSF1R than on AR (Suppl. Fig. 3). Collectively, our data demonstrate that the impact of flutamide, contingent on the expression levels of miR-449a and miR-449b-5p, is more substantial in ovarian cancer cells than in primary ovarian epithelial cells.

The involvement of the CSF1/CSF1R pathway in the progression of epithelial ovarian cancer is well-documented [17]. In addition, our own mouse models previously demonstrated the enhanced virulence associated with CSF1 overexpression in ovarian cancer and, conversely, reduced tumorigenicity in mice implanted with CSF1-negative ovarian cancer cells [18]. This result, combined with our previous observations of CSF1 and CSF1R overexpression in HR tissues [9, 10], suggested the critical role of the CSF1/CSF1R pathway in ovarian cancer initiation. Given this, our current finding that flutamide can restore the reduced expression of miR-449a and miR-449b-5p in HR tissues, thereby suppressing CSF1R, provides a mechanistic insight into how flutamide may potentially mitigate the risk of ovarian cancer initiation.

The putative role of the AR pathway in ovarian cancer development was a primary motivator for our Phase 2 clinical study, in which we tested the effects of the anti-androgen agent flutamide in HR women [9]. Our findings that flutamide effectively normalized the elevated protein biomarkers in HR patients lend further support to the important role of androgen signaling in the initiation and progression of ovarian cancer. The present study extends these findings by demonstrating that flutamide upregulates miR-449a and miR-449b-5p in SKOV3 and Hey ovarian cancer cells (Fig. 3), leading to a marked reduction of AR expression at both the mRNA and protein levels (Fig. 5 and Suppl. Fig. 5). This revelation of miR-449-mediated suppression of AR expression adds a new dimension to our understanding of flutamide action, complementing its established direct antagonistic binding to AR. That is, effectively, flutamide emerges as a ‘dual inhibitor’ of AR signaling pathways, as illustrated in Fig. 2.

The tumor-suppressive roles of miR-449a and miR-449b-5p are known in various cancers, including gastric, colon, prostate, and breast cancers [19–21]. However, their role in epithelial ovarian cancer, especially high-grade serous carcinomas, has been largely unexplored [22]. Our present study bridges this knowledge gap by showing how these miRNAs modulate the expression of CSF1R and AR in ovarian cells, implicating their significant roles in regulating CSF1/CSF1R and AR signaling pathways during the development of epithelial ovarian cancer in HR women.

Interestingly, the miR-449 family is part of the broader miR-34/miR-449 superfamily, which comprises two miRNA clusters, miR-34a/b/c and miR-449a/b/c. These clusters share a similar seed sequence and adjacent nucleotide sequences [23] and overlapping functions in mediating cell cycle arrest and apoptosis in tumor suppression [24]. Previous studies have linked the downregulation of miR-34 with ovarian cancer [25, 26]. In this study, we observed that some members of the miR-34 family followed a similar expression trend to miR-449 in ovarian tissues, with miR-34b-5p expression significantly upregulated upon flutamide treatment (Suppl. Fig. 1A, left) and miR-34c-5p/3p expression significantly upregulated in LR compared to HR subjects (Suppl. Fig. 1B, left). This trend of miR-34 expression was absent in fallopian tube samples (Suppl. Fig. 1, right), consistent with the more subdued differential expression of miR-449 in the fallopian tube compared to the ovary (Fig. 1B).

To our knowledge, this is the first study to investigate miRNAs in normal ovarian and fallopian tube tissues from women at high risk for ovarian cancer but without existing ovarian or other peritoneal cancers, tubal dysplasia, or STIC lesions. Our findings suggest that flutamide treatment can effectively restore the diminished expression of miR-449a and miR-449b-5p, among other miRNAs, in the ovarian and tubal tissues of these high-risk women. This restoration decreases the expression of critical molecular markers such as CSF1R and AR, which are associated with an elevated risk for ovarian cancer.

A well-tolerated low-dose regimen of anti-androgen flutamide [9] has been conceived as a potential chemopreventive measure for ovarian cancer. Our findings support its chemopreventive potential for high-risk women, particularly those with decreased miR-449 expression as the subgroup most likely to benefit. Further studies are needed to ascertain whether the flutamide-induced miR-449 restoration effectively translates into a reduced risk of developing ovarian cancer in these patients.

## Data Availability

All data produced in the present study are available online at the Gene Expression Omnibus (GEO) database (GSE252170).

## List of abbreviations

AR: Androgen Receptor
BMI: Body Mass Index
BRCA1/2: Breast Cancer 1/2, early onset (genes)
CSF1: Colony-Stimulating Factor 1
CSF1R: Colony-Stimulating Factor 1 Receptor
FFPE: Formalin-Fixed, Paraffin-Embedded
GAPDH: Glyceraldehyde-3-Phosphate Dehydrogenase
hEGF: human Epidermal Growth Factor
HR: High-Risk
IIT: Investigator-Initiated Trials
LR: Low-Risk
miR: microRNA
qRT-PCR: Quantitative Reverse Transcription Polymerase Chain Reaction
STIC: Serous Tubal Intraepithelial Carcinoma

## Ethics approval and consent to participate

Eligible patients were consented under the University of Arizona Institutional Review Board (IRB)-approved Flutamide trial (NCT00699907), the University of Arizona Cancer Center Tumor Biorepository (NCT00896935), and/or the HR/LR translational study (UACC IIT Continued Recruitment of High-Risk and Low-Risk Control Subjects to Assess Biomarkers of Ovarian Cancer Risk, IRB 1504771536). Together, these 3 studies comprise the Flutamide Correlative Study. These projects have been reviewed and approved by the University of Arizona IRB or designee. All documents referenced have been reviewed and approved. The University of Arizona maintains a Federal-wide Assurance (FWA) with the Office for Human Research Protections (OHRP) (FWA #00004218). This Institution assures that all of its activities related to human subjects research, regardless of the source of support, will be guided by the Belmont Report and applicable regulations according to 45 CFR 46.111 and/or 21 CFR Part 50.

## Consent for publication

(Not applicable)

## Availability of data and material

The miRNA-sequencing reads have been deposited and are available in the Gene Expression Omnibus (GEO) database (GSE252170).

## Author contributions

XW, analysis, manuscript preparation and revision; HHW, data collection, experimentation, analysis; MW, analysis; SG, data collection; MM, data collection; DR, specimen preparation, data collection; JC, resources; WZ, manuscript revision; GY, analysis, design, supervision, manuscript preparation and revision; SKC, concept, design, supervision, data collection, resources, manuscript preparation and revision. All authors reviewed and approved the manuscript. There is no conflict of interest to declare.

---

**Suppl. Figure 1.**
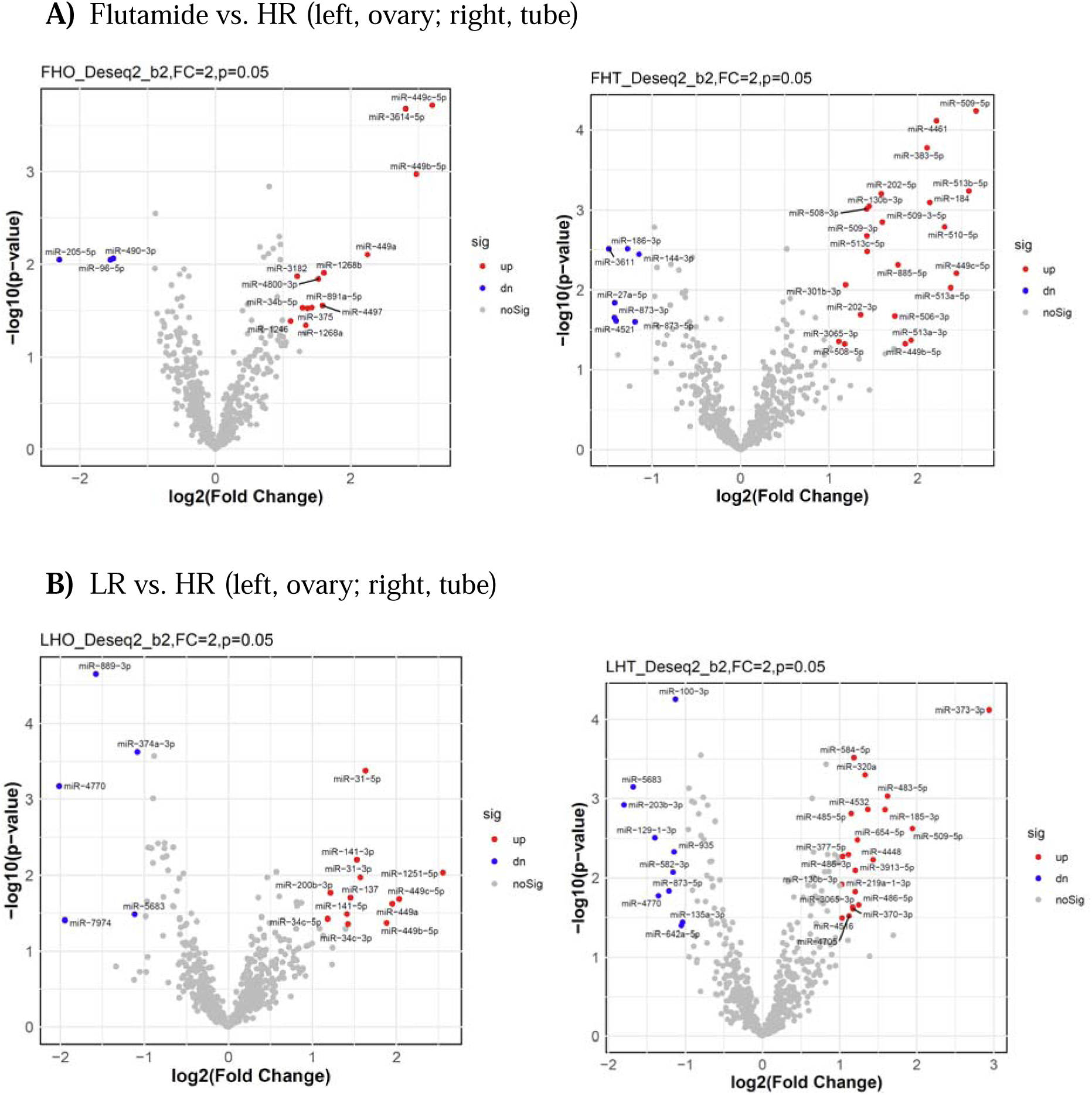
Volcano plots of miRNA expression profiles in flutamide vs. HR cohort (A) and in LR vs. HR cohort (B) from the ovary (left) and fallopian tube (right) samples. Red and blue dots indicate miRNAs that are significantly up-regulated (fold change > 2, p < 0.05) and down-regulated (fold change < 0.5, p < 0.05), respectively. The statistical significance (p-value) of the number difference between red and blue dots is 0.018 (left, A), 0.004 (right, A), 0.15 (left, B), and 0.05 (right, B), respectively.

**Suppl. Figure 2.**
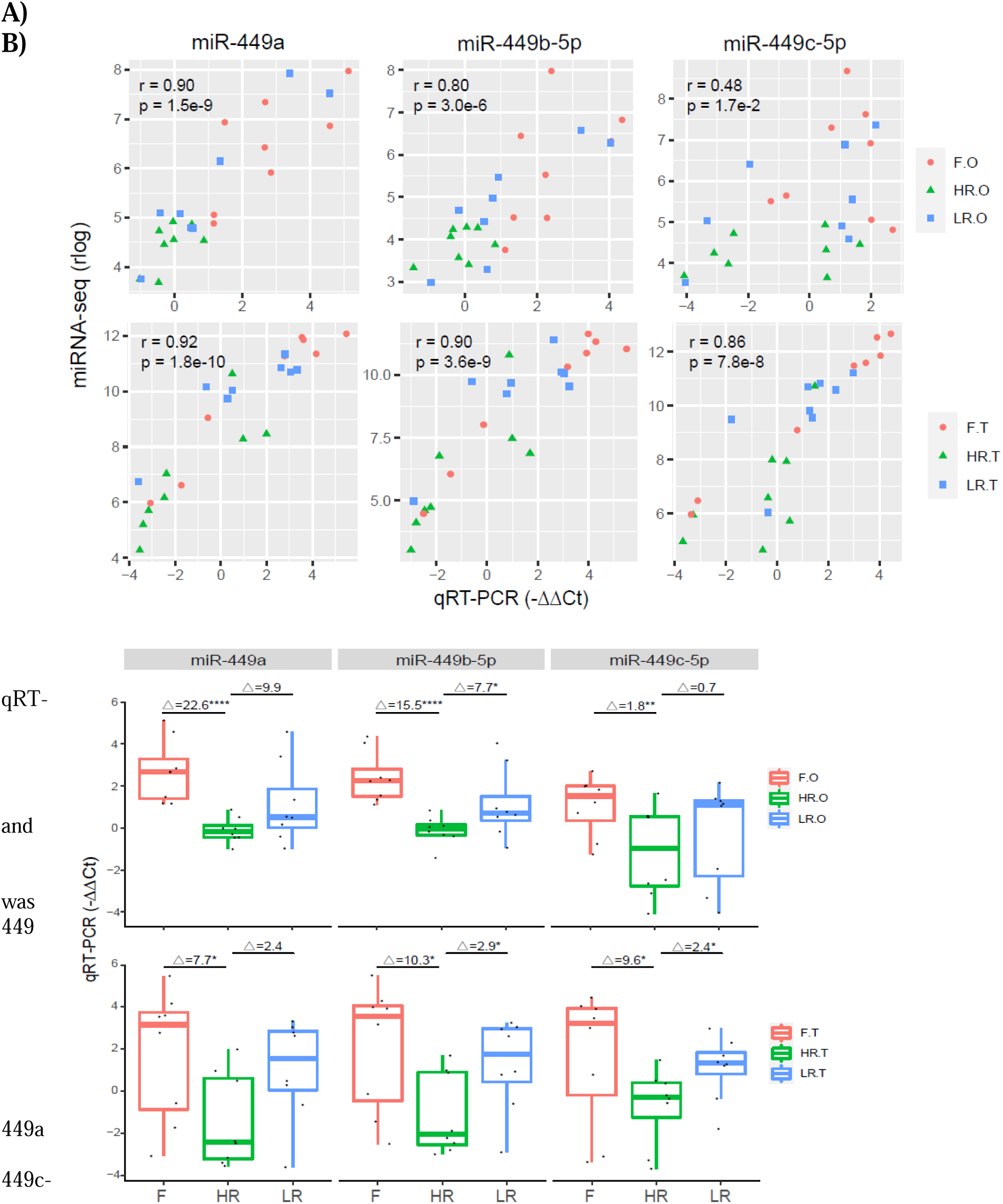
PCR Validation of miRNA-seq Results. In each patient cohort (Flutamide/F, HR, LR) from the miRNA-seq analysis, qRT-PCR used to assess miR-expression in ovarian and fallopian tube samples from eight randomly selected individuals. (A) Correlation analysis for miR-(left), miR-449b-5p (middle), and miR-5p (right) in ovarian (top) and tube (bottom) samples, between miRNA-seq results (y-axis, rlog normalized values) and qRT-PCR (x-axis, log2 of miRNA expression relative to the HR average). Each panel shows the Pearson correlation coefficient (r) and the p-value of the correlation. (B) Boxplot of miR-449 expression levels determined by qRT-PCR (y-axis, log2 of miRNA expression relative to the HR average) in ovarian (top) and tube (bottom) samples. Glass’s delta effect size and p-values (*, **, and **** indicating p < 0.05, 0.01, and 0.0001, respectively) for each pairwise comparison are shown at the top of each boxplot.

**Suppl. Figure 3.**
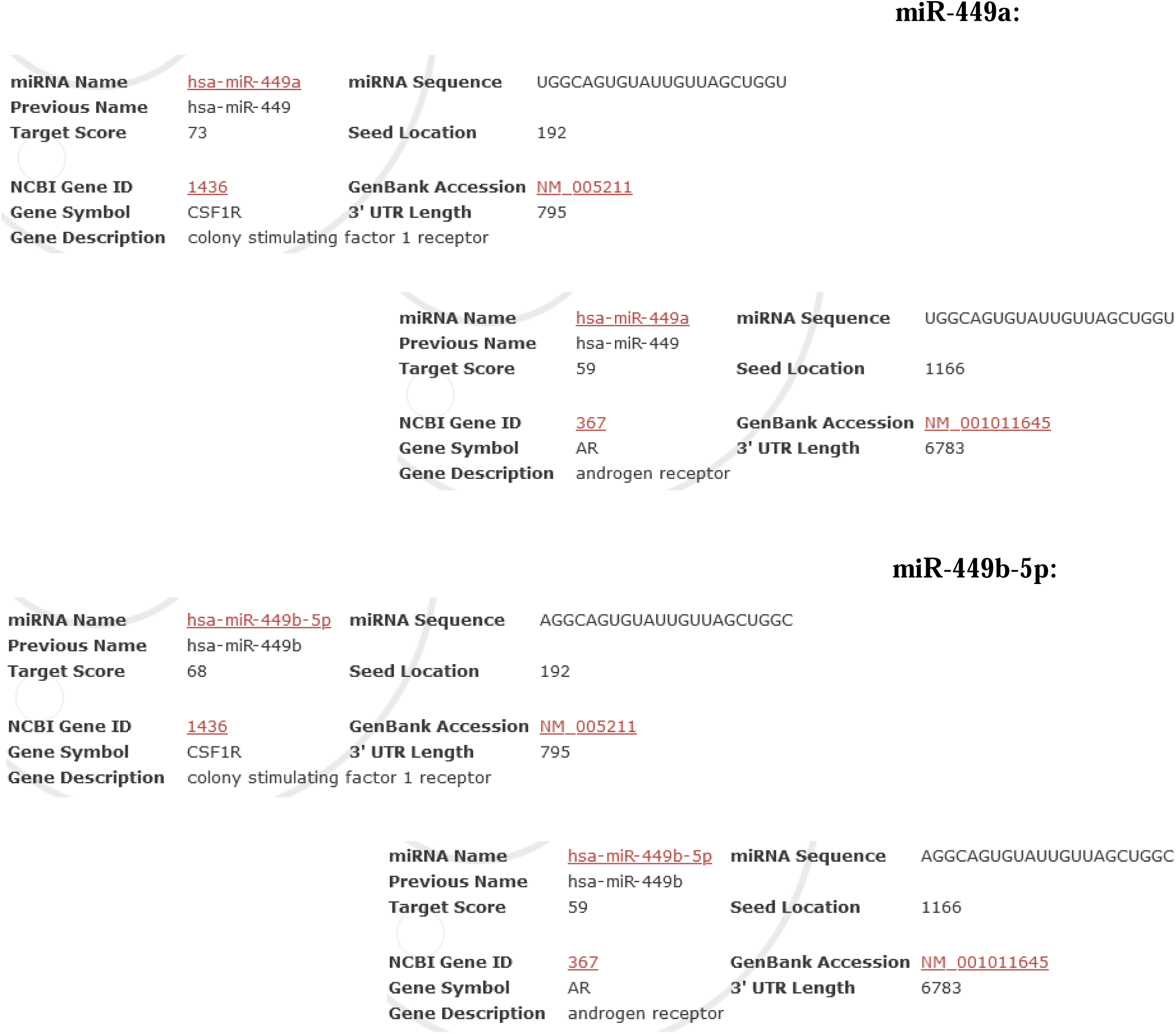
Computational prediction of miR-449a and miR-449b-5p targeting CSF1R and AR. The predictions were obtained from miRDB, an online database for miRNA target predictions and functional annotations [16].

**Suppl. Figure 4.**
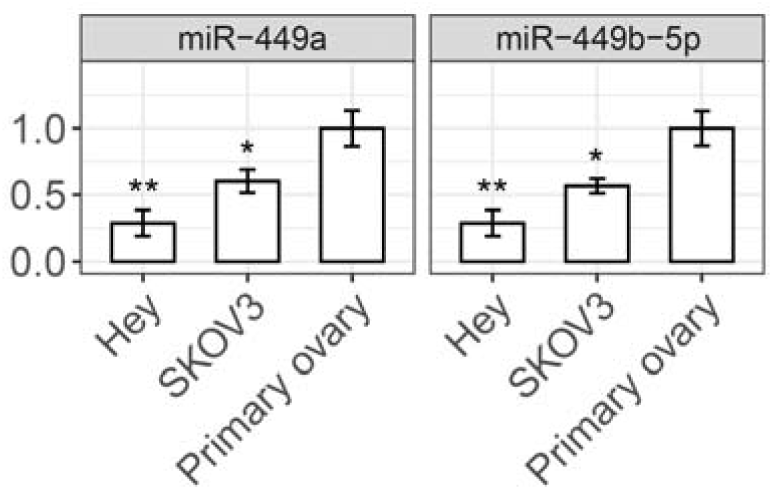
The expression levels of miR-449a and miR-449b-5p in Hey and SKOV3 ovarian cancer cells and primary ovarian epithelial cells, measured by qRT-PCR. The statistical significance (if any) of each pairwise comparison to the primary ovarian cell control is indicated with * and ** for p < 0.05 and 0.01, respectively. Error bar, s.e.m.

**Suppl. Figure 5.**
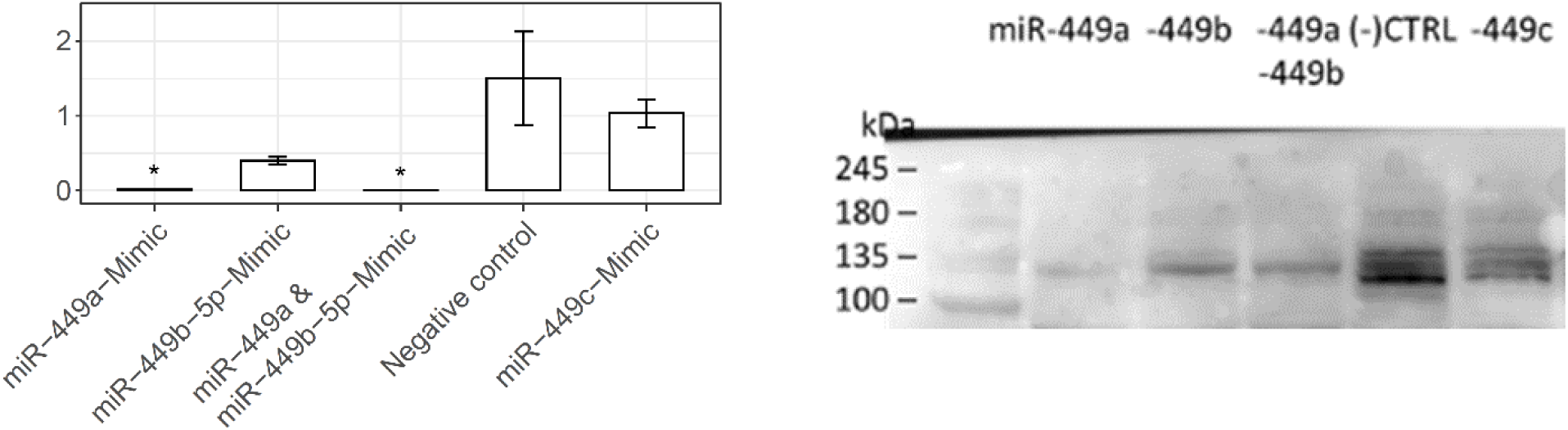
CSF1R mRNA and protein in Hey cells with miR-449 mimics. (Left) CSF1R mRNA levels at indicated conditions, measured by qRT-PCR (n=3). The statistical significance (if any) of each pairwise comparison to the negative control is indicated with * for p < 0.05. Error bar, s.e.m. (Right) CSF1R protein levels at indicated conditions measured by Western blot. Dual mimics (miR-449a and miR-449b-5p) showed a similar effect as a single miR-449a mimic on CSF1R mRNA and an intermediate effect between miR-449a and miR-449b-5p on CSF1R protein. miR-449c-5p had little effect on CSF1R mRNA and protein.

**Suppl. Figure 6.**
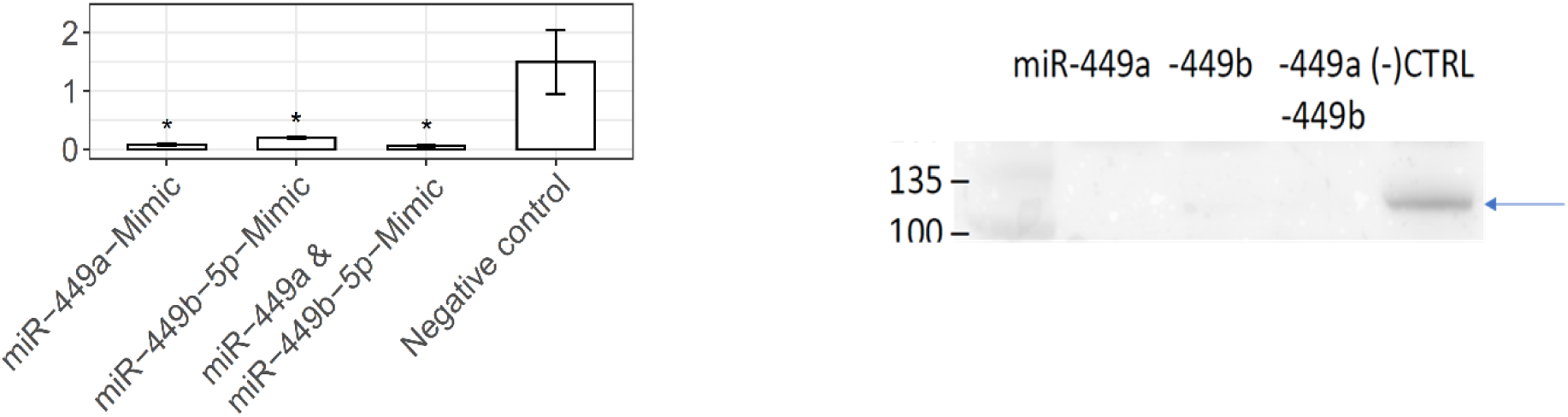
AR mRNA and protein in Hey cells with miR-449a and miR-449b-5p mimics. (Left) AR mRNA levels at indicated conditions, measured by qRT-PCR (n=3) The statistical significance of each pairwise comparison to the negative control is indicated with * for p < 0.05. Error bar, s.e.m. (Right) AR protein levels at indicated conditions, measured by Western blot.

**Suppl. Figure 7.**
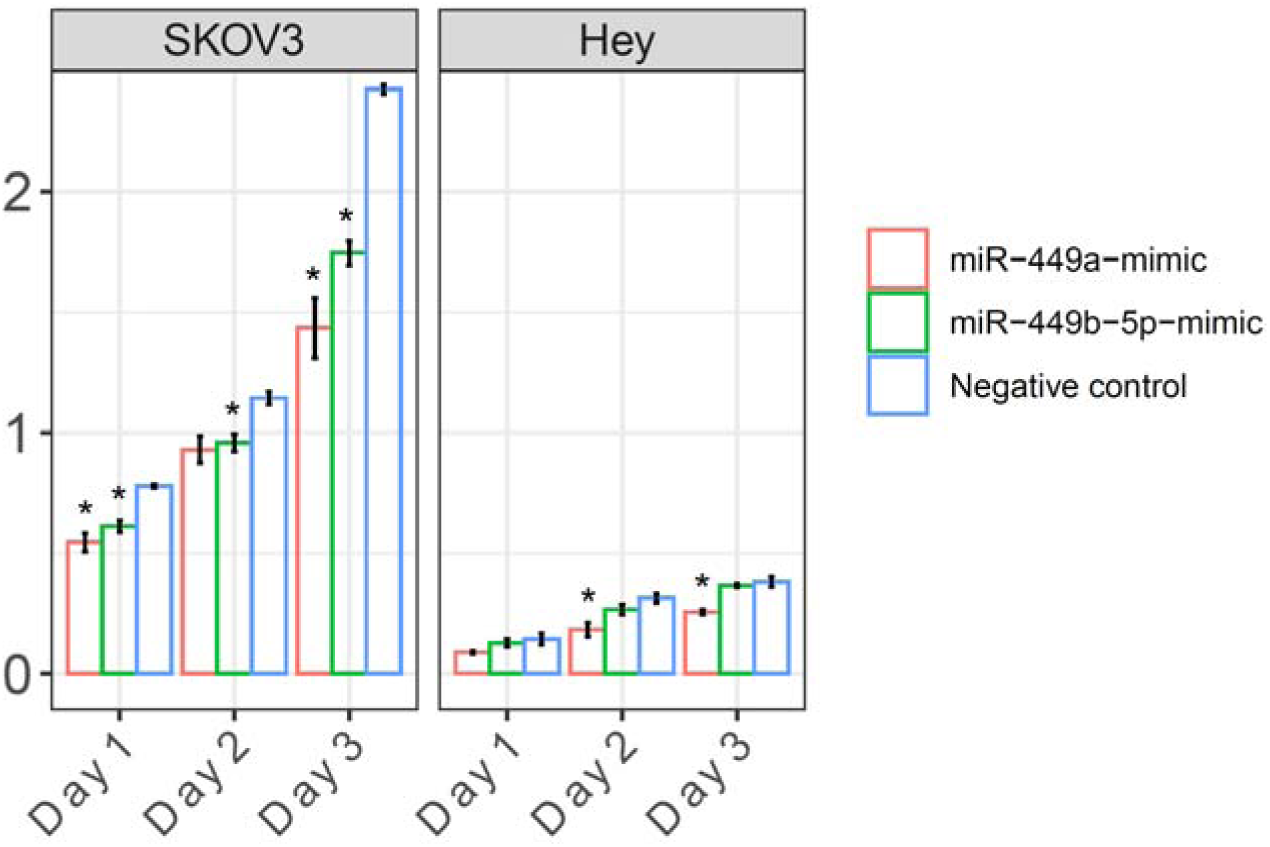
miR-449a and miR-449b-5p mimics inhibit the proliferation of ovarian cancer cells (SKOV3, left; Hey, right). After transient transfection of mimics or negative control as described in Methods, cells were seeded and then followed for 3 days for cell confluency change (n=2, see Methods for detail). The statistical significance (if any) of each pairwise comparison to the negative control is indicated with * for p < 0.05. Error bar, s.e.m.

